# Harmonization of cerebral blood flow measurements by multi-delay 3D gradient and spin echo, and single-delay 2D echo planar imaging

**DOI:** 10.1101/2025.06.20.25328792

**Authors:** Josiah B. Lewis, Chunwei Ying, Michael M. Binkley, Melanie E. Fields, Igor Dedkov, Slim Fellah, Jingyi Zhang, Amy Mirro, Joshua S. Shimony, Yasheng Chen, Jin-Moo Lee, Andria L. Ford, Hongyu An, Kristin P. Guilliams

## Abstract

**Purpose:** Cerebral blood flow (CBF) is commonly measured by pseudo-continuous arterial spin labeling (PCASL) in human research, but recent advancements in methodology have limited data reuse. The object of this work is to harmonize two distinct PCASL techniques within a cohort with a wide range of CBF values.

**Methods:** Participants had two PCASL sequences collected within a single session: a single post-label delay sequence with a 2D echo-planar imaging (EPI) readout, “CBF_2D,1PLD_”, and a five post-label delay sequence with gradient and spin echo (GRASE) 3D readout, “CBF_3D,5PLD_”. Linear regression modeling to impute CBF_3D,5PLD_ from CBF_2D,1PLD_, hemoglobin, and age were assessed within gray matter (GM) and white matter (WM) using leave-one-out cross-validation for prediction errors and confidence intervals. Within-subject coefficient of variation (wsCV) and inter-class correlation coefficient (ICC) were calculated using CBF_3D,5PLD_ imputed vs. measured as pseudo test-retest pairs.

**Results:** Fifty participants, ages 8–45 (median 25) years, had usable CBF_3D,5PLD_ and CBF_2D,1PLD_, including 17 participants with sickle cell disease (SCD), who were matched by age (*p* = 0.90) and sex (*p* = 0.16) to those without SCD. A multiple linear regression model including hemoglobin and age fit GM CBF (R^2^_adj._ = 0.82; for WM CBF R^2^_adj._ = 0.78). The wsCV for CBF_3D,5PLD_ was 9.1% for GM, 11.3% for WM. ICC was 0.89 for GM and 0.87 for WM. Models without age or hemoglobin fit slightly worse.

**Conclusion:** Our study demonstrates feasibility to impute 3D-GRASE multi-PLD CBF from a 2D-EPI single-PLD technique, which promotes data sharing and harmonization.

## 1. Introduction

Pseudo-continuous arterial spin labeling (PCASL) magnetic resonance imaging (MRI) is used broadly to measure cerebral blood flow (CBF) for research and clinical applications. It has the advantages of not requiring ionizing radiation, injected contrast, or arterial blood sampling, making it an attractive perfusion measurement technique for human subjects, particularly children, compared to alternatives such as dynamic susceptibility contrast (DSC) MRI and positron emission tomography (PET). However, adoption of quantitative CBF by ASL has been difficult to achieve, as demonstrated by test-retest variability [1,2], and variability due to different spin-labeling techniques [1,2] and scanner models [3].

Different implementations of PCASL CBF have been unavoidable, including variation in hardware and firmware, as well as institutional and lab-specific differences in MRI CBF collection and processing. With the adoption of PCASL sequences and processing procedures with improved resolution, higher SNR, shorter acquisition time, and technique adjustments to align with consensus, data laboriously collected using prior techniques may be under-utilized due to unknown compatibility with data from other techniques, including from those that replaced them as the standard for research. This diminishes the statistical power and diversity of MRI CBF datasets, which is deleterious in human research, especially pediatric, longitudinal, and rare disease studies, as well as whole-brain correlation studies, which suffer in low-N scenarios [4]. Many research studies to date have abided by principles and clinical recommendations laid out by Alsop et al., 2015, which were based on a single post-label delay (PLD) and 2-dimensional echo planar imaging (2D-EPI) or alternatively 3D gradient and spin echo (3D-GRASE) readout [5]. Multi-PLD techniques have increasingly replaced single-PLD sequences for some research applications, largely due to their ability to correct for delayed or variable transit times by computing per-voxel arterial transit time (ATT), but involve more complex sequence design, can take longer to acquire, and can be more complex to process and interpret [6–8]. 3D GRASE readout methods with background correction have increasingly replaced other, usually lower-resolution readouts, and were recommended for clinical applications by a recent consensus work, Lindner et al. 2023, with 2D-EPI readout as an alternative [9]. In addition to differences in background correction, signal strength, noise inherent in the different readouts, and resulting differences in optimal processing steps, ATT is an unmeasured confounder of the single-PLD calculation of CBF, whereas for the multi-PLD data, ATT is modeled and included in the CBF calculation. As a result, there are multiple potential sources of disagreement in CBF calculated using 2D-EPI vs. 3D-GRASE PCASL readouts [9,10] and single-PLD vs. multi-PLD [7,11], precluding a simple adjustment factor from one methodology to another.

We sought to harmonize CBF values in GM and WM from one PCASL technique with another, by creating and evaluating regression models between a lower-resolution 2D-EPI readout, single-PLD (1PLD) sequence and a higher resolution 3D-GRASE, five-PLD (5PLD) readout sequence that replaced it in research use at our institution, using paired data from a retrospective cohort with an expected wide range of CBF values.

## 2. Methods

### 2.1 Study Design

All data were collected at Washington University in Saint Louis School of Medicine with institutional review board approval and informed consent from participants or their parents/legal guardians. This study used retrospectively collected data from research visits of children and adults 8–45 years old. Paired data was collected from two different PCASL sequences during a period of transition at our institution from one PCASL CBF technique to another. To understand whether harmonization of the two CBF techniques was equally valid across a range of CBF values, and to understand the role of hemoglobin (hb) value in harmonization, we included study participants with and without sickle cell disease (SCD). Therefore, our models are based on an example of real-world research data collected using a lower-resolution, less cutting-edge technique, data which can only be reused alongside higher-resolution, more cutting-edge data if the CBF generated by the two techniques can be harmonized. A blood draw was completed during the study visit to measure hemoglobin. Participant demographics including age and sex were also collected.

### 2.2 Magnetic resonance imaging

#### 2.2.1 Overview of MRI

MRIs were conducted on one of two Siemens Prisma 3T MRI scanners with a Syngo version of VE11C, and a 32- or 64-channel head coil. Two different PCASL sequences for CBF calculation were collected during the same MRI session, along with T1-weighted magnetization prepared rapid gradient echo (MPRAGE) sequence for tissue segmentation, a FLAIR image for lesion assessment and masking, and a fast inversion recovery sequence (FASTIR) [12] to image the superior sagittal sinus for calculation of the longitudinal relaxation time for blood (T1_blood_).

#### 2.2.2 PCASL sequences

Key features of the two PCASL sequences are presented here and summarized in Table 1. The first PCASL sequence uses a 2D-EPI readout and 1PLD (labeling plane 90 mm below the middle slice, TE / TR = 12 ms / 3810 ms, flip angle = 90°, echo spacing = 0.49 ms, number of slices = 18, slice thickness = 5 mm with 1 mm gap, slice oversampling was off, in-plane voxel dimensions = 3.4 × 3.4 mm, FoV = 220 mm, phase partial Fourier = 6/8, bandwidth = 2264 Hz/Px). Forty label/control pairs were collected. The nominal post-label delay is 1.5 s, as recommended for pediatric studies [5]. The actual delay from labeling to readout is different for each slice, with slice acquisition times equal to approximately PLD + 42 ms per slice, starting with the most inferior slice and moving axially in the superior direction, such that the first slice has a PLD of 1.5 s, and the 18^th^ and final slice has a PLD of approximately 2.2 s. Acquisition time was 5 min 9 s. The young adults included in this study were being scanned in a predominantly pediatric cohort, so all participants, including adults, were scanned with PLD = 1.5 s for consistency.

**Table 1.** Comparison of two pseudocontinuous arterial spin labeling (PCASL) techniques for cerebral blood flow measurement.

|  | **CBF 2D 1PLD** | **CBF 3D 5PLD** | **Note** |
| --- | --- | --- | --- |
| *Acquisition* |  |  |  |
| PLD (s) | 1.5 | 0.5, 1, 1.5, 2, 2.5 |  |
| Label/Control Pairs | 40 | 2, 2, 2, 3, 3 |  |
| Resolution x, y, z (mm) | (3.4, 3.4, 5) | (2.5, 2.5, 2.5)* | *Point spread function may be larger |
| Readout | 2D-EPI | 3D-GRASE |  |
| Acquisition Time | 5:09 | 7:32 |  |
| Background Suppression | No | Yes |  |
| *Processing Steps* |  |  |  |
| Motion Correction | Yes | No |  |
| Model | One-compartment | One-compartment |  |
| T<sub>1blood</sub> | Measured at visit | Measured at visit | Hemoglobin modeled in 7 cases |
| $\alpha$ (labeling efficiency) | 0.85 | 0.77 = 0.85*0.95*0.95 | |
| $\lambda$ (blood/tissue partition) | 0.9 | 0.9 | |
| ATT calculated | No | Yes |  |

The second PCASL sequence uses a 3D-GRASE readout with 5PLD (labeling plane 90 mm below the center of the image volume, TE / TR = 23.38 ms / 4300 ms, Flip angle = 120°, turbo factor = 28, echo spacing = 0.53 ms, segments = 4 (2 in ky, 2 in kz), voxel dimensions = 2.5 × 2.5 × 2.5, slice oversampling = 10%, phase partial Fourier was off, bandwidth = 2264 Hz/Px). Background suppression was employed for label/control images. The five post-label delays and corresponding number of control/label pairs collected for each PLD were 0.5, 1.0, 1.5, 2.0, and 2.5 seconds, and 2, 2, 2, 3, and 3 pairs, respectively. A first frame (M_0_) image was collected without background suppression. Acquisition time was 7 min 32 s.

#### 2.2.3 Other MRI sequences

The skullstripped T1-weighted MPRAGE image was segmented into GM and WM probability maps calculated by the segmentation tool SPM [13]. Lesions were manually delineated on native FLAIR images and aligned to the T1-weighted image using FMRIB’s Linear Registration Tool (FLIRT) [14].

### 2.3 CBF quantification

#### 2.3.1 Unified analysis steps

Key features of quantification of the two CBF values are presented here and summarized in Table 1. A skullstrip was computed on the first frame of each sequence using the Brain Extraction Tool (BET) [15]. If BET did not yield a sufficiently high quality skullstrip, the Brain Surface Extractor (BSE) [16] was used. If both methods failed, the participant was excluded. A nonlinear warp was computed from MPRAGE to each of the two PCASL spaces using the skullstripped MPRAGE and the M_0_ of the skullstripped, upsampled PCASL, using Advanced Normalization Tools (ANTS) [17]. This warp, followed by a downsampling step, was then used to transform the MPRAGE GM and WM probability images and FLAIR lesions masks from MPRAGE space into PCASL space.

#### 2.3.2 CBF 2D, 1PLD

For the 2D-EPI 1PLD sequence, the frames were corrected for motion using the affine registration tool McFlirt, with normalized correlation as the cost function and four-stage alignment [14]. The images were smoothed using a truncated Gaussian filter with sigma = 0.4 voxels.

Mean label and control images were created and partial volume corrected for GM and WM based on linear regression to the near neighbor voxels [18]. We defined the neighborhood for partial volume correction as ± 2, 2, 1 voxels in x, y, z, directions i.e., ± 6.8, 6.8, 5 mm.

CBF_2D,1PLD_ was calculated using the following one-compartment model [19], since this model is most commonly recommended for research, e.g., by Alsop et al. [5]:

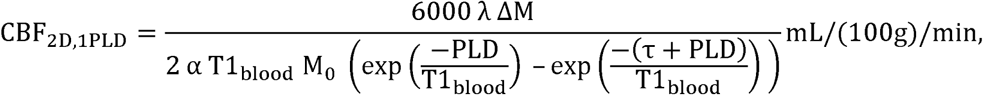

where the brain/blood partition coefficient λ = 0.9 g/mL, the labelling efficiency α = 0.85 [5]. T1_blood_ is the longitudinal relaxation time of blood, measured in milliseconds, which is calculated from an ROI in the superior sagittal sinus imaged by a sequence or inferred from the hb using the model T1_blood_ = -41.3×hb + 2230. This model is generated using linear regression of inversion recovery measurements paired with hb values from other research studies at Washington University and is part of our standard processing. τ is the labeling train duration. The average control image is used as M_0_. The difference between time average label and control image intensities, ΔM, is filtered such that ΔM < 0 and M_0_ < 200 are excluded to avoid physiologically impossible results in CBF maps.

#### 2.3.3 CBF 3D, 5PLD

The 3D-GRASE 5PLD images were visually screened for motion and those with obvious motion were excluded. Background suppression in this sequence minimizes the structural signal collected in the label and control images, and the contrast changes significantly with each PLD, making it non-optimal for motion correction. The M_0_ was filtered to remove voxels with maximum possible values, indicative of artifact. Subsequent partial volume correction effectively smooths the images, so we did not smooth the PCASL signal at this step in the processing.

Mean label and control images were created for each PLD and underwent partial volume correction for GM and WM based on linear regression to the near neighbor voxels. The M_0_ was also partial volume corrected. For 5PLD PCASL the neighborhood is selected to approximate the same size as the 1PLD technique as close as possible given different voxel sizes, ± 3, 3, 2 voxels, that is, ± 7.5, 7.5, 5 mm.

CBF_3D,5PLD_ for each of the PLDs is calculated using the same one-compartment model as CBF_2D,1PLD_, except the calculated ATT is used if PLD < ATT, and PLD is used when PLD > ATT:

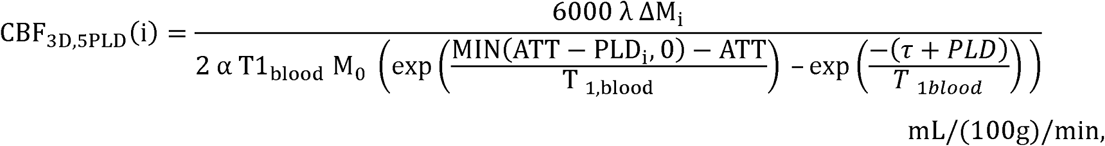

where ATT is calculated from the weighted delay, *WD*, through a monotonic function, using a lookup table [7,20]:

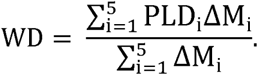

The same value is used for the brain/blood partition coefficient λ = 0.9 g/mL as was used for CBF_2D,1PLD_, but the labelling efficiency α = 0.85 × 0.95 × 0.95 ≈ 0.77 for CBF_3D,5PLD_, where a base efficiency of 0.85 is modified by a factor of 0.95 for each of two background suppression pulses. CBF_3D,5PLD_ is calculated as the mean of the CBF_3D,5PLD_(i) measurements. This model reduces to the CBF_2D,1PLD_ model in the case PLD > ATT.

### 2.4 ATT > PLD analysis

If ATT > PLD for a voxel, the blood will not have arrived at the local tissue by the time of readout, and CBF_2D,1PLD_ will be underestimated. For our young adult participants, the conventional PLD is 1.8 s rather than 1.5 s [5]; however, to combine with pediatric data we used the 1.5 s recommended for the young population for all our participants. We are able to check the validity of this comparison as we also calculate ATT. It should be noted that per-slice effective PLD increases axially for our CBF_2D,1PLD_ sequence. We calculated maps of ATT – PLD_effective_, the difference between the modeled ATT and the effective PLD for a given voxel, and calculated the median, IQR, and max % of voxels where ATT > PLD. We co-registered the ATT – PLD_effective_ maps to the MNI ICBM 152 atlas [21] to create a map of which voxels and regions most often experienced ATT > PLD_effective_.

### 2.5 Statistical analyses

We used Wilcoxon matched-pairs signed rank test to test for significant differences between CBF_2D,1PLD_ and CBF_3D,5PLD_. The Pearson’s correlation coefficient was used to perform univariate comparisons of continuous variables, including CBF, ATT, hb, and age. Baseline demographics in continuous variables are reported as median and inter-quartile range (IQR, 25% and 75%). To test for differences between participants with and without SCD, we used the Mann-Whitney *U* test for age, and the chi-squared test for sex. The criterion for significance was defined as a *p* value of <0.05, and we report confidence intervals (CIs) of 95%.

Least-squares single and multiple linear regression fits were used to model the relationship between CBF calculated from the two MRI sequences and generate an adjusted R^2^ (R^2^_adj._) statistic. We confirmed that CBF_2D,1PLD_, age, and hb were not multicollinear using the variance inflation factor. To assess the models, we calculated the Akaike information criterion (AIC) and Bayesian information criterion (BIC). We used analysis of variance (ANOVA) to test whether adding variables to simpler models significantly improved performance (*F*-statistic test). To compare the variability of our imputation technique with inherent variability in PCASL measurement, measured in test-retest studies, we used measured and imputed CBF_3D,5PLD_ as pseudo test-retest data to calculate the within-subject coefficient of variation (wsCV) [22]. We also calculated the inter-class correlation coefficient (ICC) [23], using a single measurement, absolute-agreement, 2-way mixed-effects model form [24]. Testing a model on the fitting data may overestimate how well the model will perform when applied to new data due to non-representative data and instability due to covariation, which we expect in CBF, ATT, age, and hb. So we used leave-one-out cross-validation [25] to calculate the prediction error, CI, and residual sum of squares (RSS). See Appendix Afor additional details.

## 3. Results

### 3.1 Cohort

Paired 2D-EPI 1PLD and 3D-GRASE 5PLD PCASL data were collected from 60 participants. Upon CBF processing the results for two adult controls were found to have been corrupted by a high degree of motion and were therefore excluded. Eight participants were excluded due to a lack of hb measurement contemporaneous with the MRI scan. The final cohort used for analysis consisted of 50 participants, median age 22.5 years old (IQR 16–27 years, full range 8–45) (Table 2). Twenty were male, 30 were female. Median hb was 11.8 g/dL (IQR 10.0–13.1). Forty-six scans were acquired on one Siemens Prisma 3T scanner, 4 on another. In the final cohort, 17 participants had anemia from SCD; the remaining 33 participants did not. There was no difference in median age (*p* = 0.90) or sex (*p* = 0.37) between those with and without SCD. For seven of the participants (three with SCD), the fast inversion recovery image of the superior sagittal sinus was not collected or was of inadequate quality. In these seven cases T1_blood_ was inferred from a hb model, such that CBF was calculated with a near-linear dependence on hb (via T1_blood_) for these participants (Figure S1).

**Table 2.**
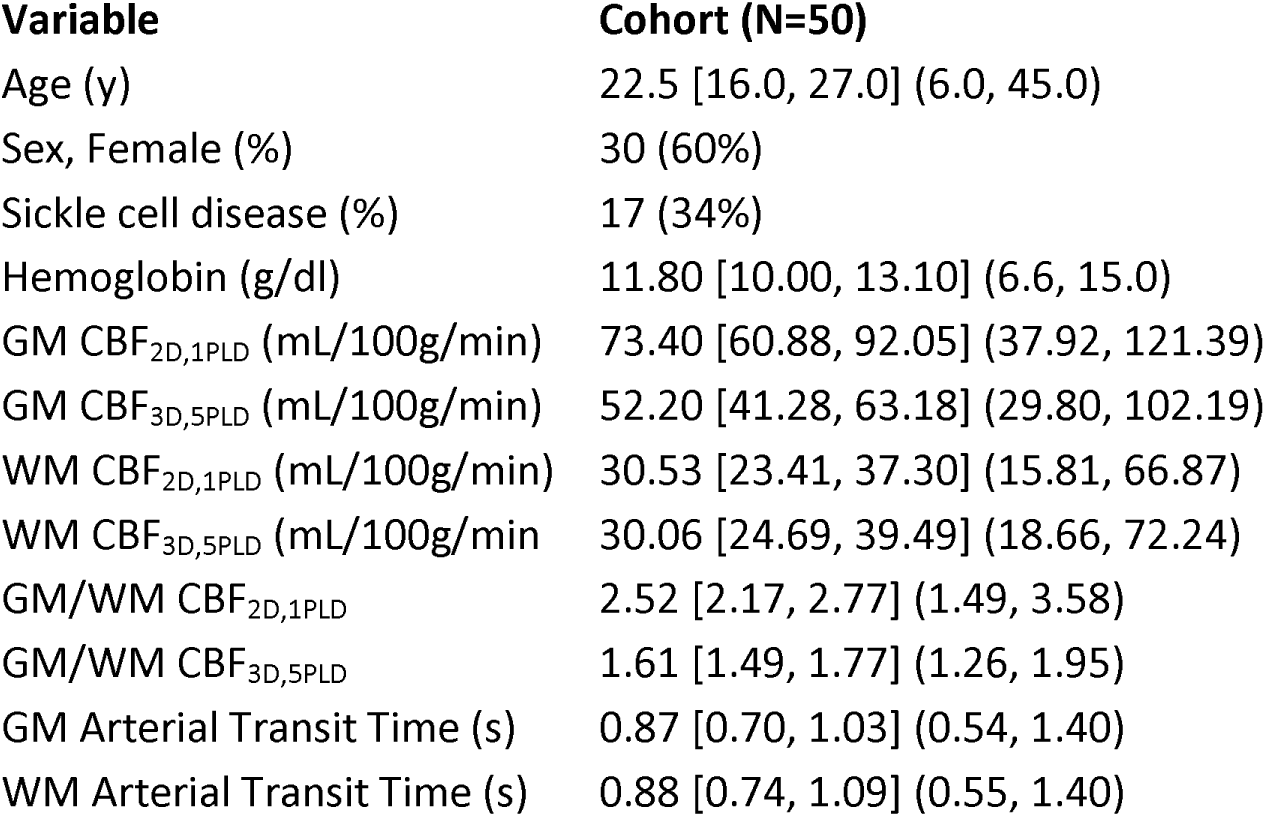
Demographics and baseline values of cohort presented as Median [interquartile range] (minimum, maximum), except for sex, and sickle cell disease, which are reported as number (percentage).

### 3.2 Models of gray matter CBF

In GM, CBF_2D,1PLD_ was higher than CBF_3D,5PLD_ (*p* < 0.001), and had greater range (Figure 1(a), Table 2, GM CBF_2D,1PLD_ = 73.40 [60.88, 92.05] mL/100g/min vs. GM CBF_3D,5PLD_ = 52.2 [41.28, 63.18] mL/100g/min). A Bland-Altman plot (Figure 1(b)) highlights that the difference between the two measures can be characterized as a fixed bias in the GM CBF_3D,5PLD_ of -37%±12%, CI = [-61, -13] %. There was non-significant proportional bias (*p* = 0.11, slope = 0.10 ΔCBF% /

**Figure 1.**
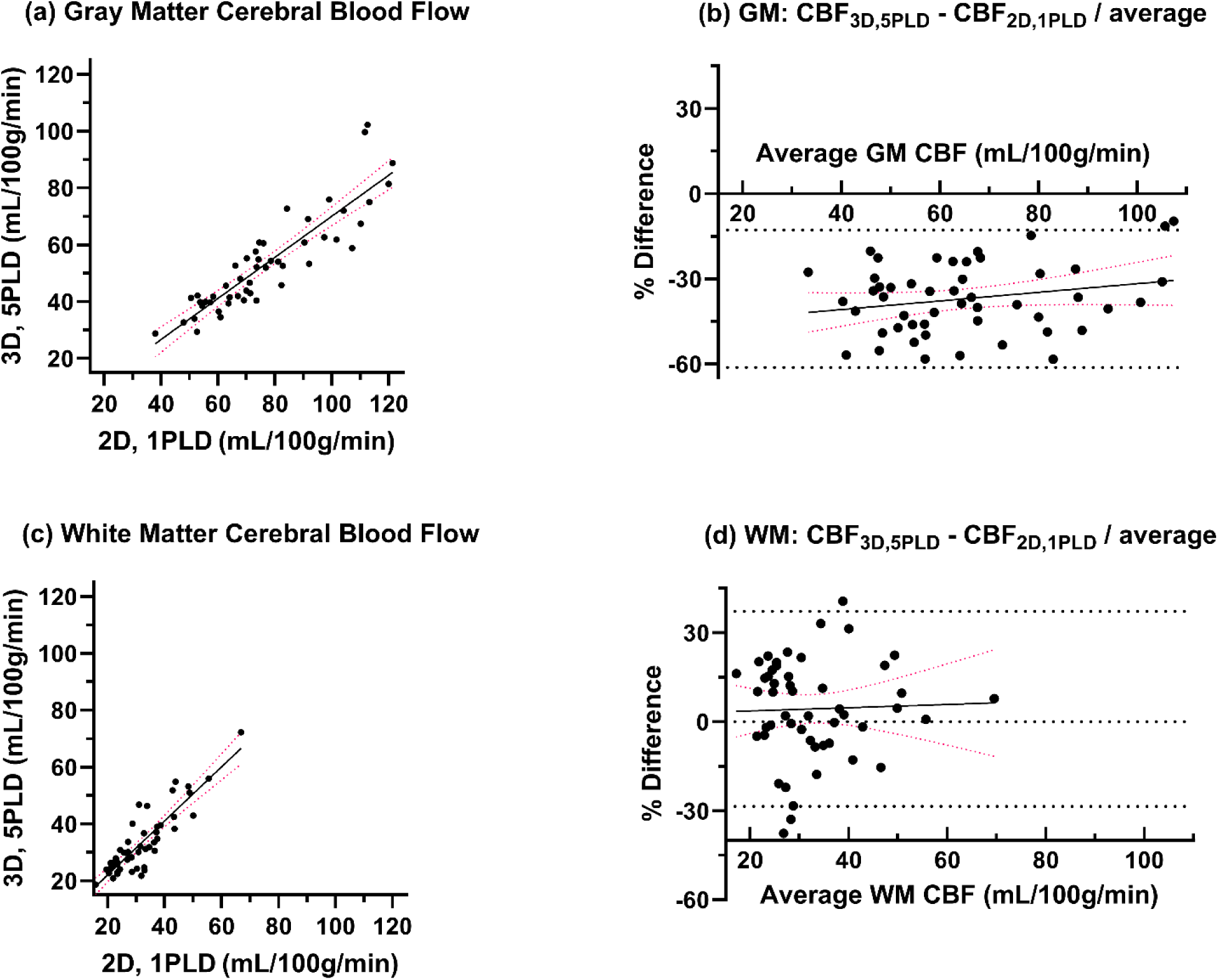
Comparison of CBF from 2D-EPI 1PLD PCASL and 3D-GRASE 5PLD PCASL techniques for GM (a, b) and WM (c, d), showing (a, c) the scatter with trend line (GM: ρ = 0.89, *p* < 0.001; WM: ρ = 0.88, p < 0.001) and 95% confidence intervals, and (b, d) Bland-Altman method comparison (GM: fixed bias = -37%±12%, CI = [-61, -13] %; WM: fixed bias = 4%±17%, CI = [-29, 37] %) with linear fit demonstrating no significant proportional bias (GM: *p* = 0.11, WM: *p* = 0.82).

CBF_mean_, CI = [-0.04, 0.34]. We fit a simple linear regression model to predict voxel-wise mean CBF_3D,5PLD_ from CBF_2D,1PLD_ in GM: *CBF_3D,5PLD_(GM) = 0.763×CBF_2D,1PLD_(GM) - 0.465 [ mL/100g/min],* with R^2^_adj._ = 0.78 and *p* < 0.001 (Figure 1(a), Table 3).

**Table 3.** Gray and white matter CBF_3D,5PLD_ imputation linear regression models and performance.

| <sup>a</sup>Model | CBF 2D,<br>1PLD | (CBF 2D,<br>1PLD)<sup>2</sup> | Hemoglobin | Age | Intercept | R<sup>2</sup><sub>adj.</sub> | σ [CI] (%)<sup>c</sup> | wsCV<sup>d</sup> | ICC<sup>d</sup> | AIC<sup>d</sup> | BIC<sup>d</sup> | RSS<sup>d</sup> |
| --- | --- | --- | --- | --- | --- | --- | --- | --- | --- | --- | --- | --- |
| *Gray Matter* |  |  |  |  |  |  |  |  |  |  |  |  |
| CBF<sub>2D,1PLD</sub>, hb, age | 0.81 (<0.001) | -- | 1.47 (0.008) | 0.28 (0.04) | -31.46 | 0.82 | 11.91 [-23.11, 23.57] | 9.1 | 0.89 | 345 | 355 | 2932 |
| CBF<sub>2D,1PLD</sub> | 0.71 (<0.001) | -- | -- | -- | -0.47 | 0.78 | 12.46 [-23.00, 25.86] | 9.6 | 0.87 | 353 | 359 | 3328 |
| *White Matter* |  |  |  |  |  |  |  |  |  |  |  |  |
| CBF<sub>1PLD</sub>, hb, age | 1.02 (<0.001) | -- | 0.87 (0.03) | -0.08 (0.36) | -7.24 | 0.78 | 17.15 [-28.17, 39.06] | 11.3 | 0.87 | 315 | 324 | 1501 |
| CBF<sub>1PLD</sub> | 0.95 (<0.001) | -- | -- | -- | 3.06 | 0.77 | 17.56 [-24.74, 44.08] | 11.6 | 0.87 | 316 | 322 | 1552 |

<sup>a</sup>Independent variable fits are presented as *coefficient (p value)*. Models were fit with data in the following units: CBF: *mL/100g/min*, hemoglobin: *g/dl*, age: *year*.
<sup>b</sup>Hemoglobin is dropped from the non-SCD only model because it is insignificant, *p* = 0.54 for GM, *p* = 0.68 for WM.
<sup>c</sup>σ is % prediction error, the standard deviation of the leave-one-out cross-validation prediction errors, with 95% confidence interval.
<sup>d</sup>Within-subject coefficient of variation (wsCV, lower is better), inter-class correlation (ICC, higher is better), Akaike information criterion (AIC, lower is better), and Bayesian information criterion (BIC, lower is better) are calculated using the base models. For residual sum of squares (RSS, lower is better), we use the prediction values from the leave-one-out cross-validation. AIC and BIC cannot be used to compare models fit using different numbers of participants.

CBF is known to vary with both hb and over the pediatric–young-adult age range. ANOVA showed significant improvement over the model with CBF_2D,1PLD_ alone after adding hb and age (*p* = 0.007),:

*CBF_3D,5PLD_(GM) = 0.809×CBF_2D,1PLD_(GM) + 1.46×hb + 0.281×age – 31.46* [mL/100g/min], with R^2^_adj._ = 0.82 (Figure 2(a), Table 3). wsCV = 9.1% and ICC = 0.89.

**Figure 2.**
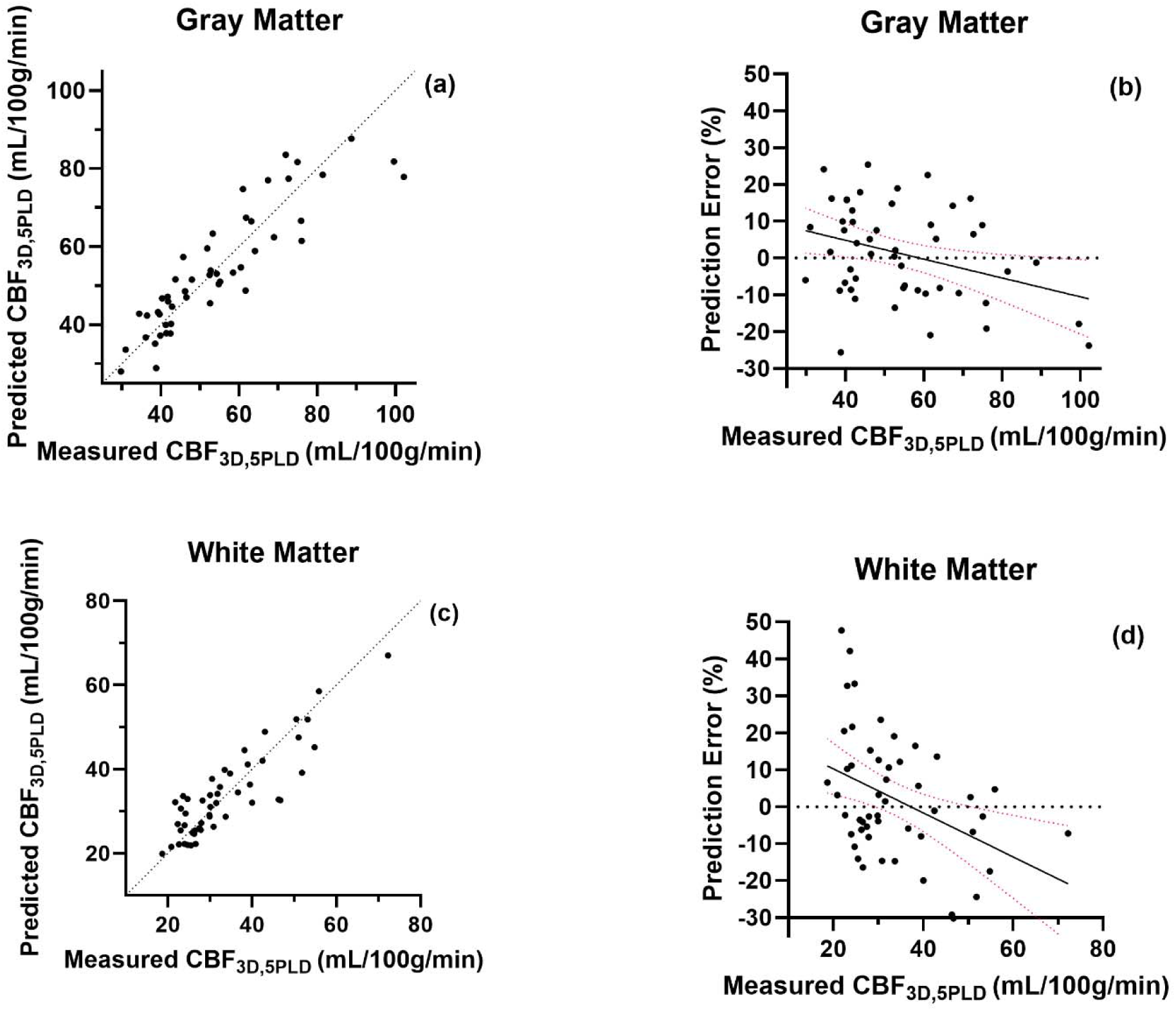
Performance of the linear regression model CBF_3D,5PLD_ ∼ (CBF_2D,1PLD_, hemoglobin, age) for GM (a, b) and WM (c, d), showing (a, c) the scatter of measured and predicted CBF_3D,5PLD_ using leave-one-out cross-validation, with a line of identity, and (b, d) % prediction error vs. measured CBF_3D,5PLD_ (GM: error = 11.91%, CI = [-23.11, 23.57] %; WM: error = 17.15%, CI = [-28.17, 39.06] %) with linear fits demonstrating significant negative trends in % error with higher CBF (GM (b): slope = -0.26%/mL/100g/min CI = [-0.46, -0.049], *p* = 0.016, R^2^ = 0.11; WM: slope = -0.60%/mL/100g/min CI = [-0.99, -0.21], *p* = 0.046 R^2^ = 0.16.

This model has lower AIC and BIC compared to the model with just CBF_2D,1PLD_. Comparing the leave-one-out cross-validation predictions to measured, we found this model has error of ±11.91% (CI = [-23.11, 23.57], Figure 2(b)) for hb, age, and CBF_2D,1PLD_ within the ranges included in our study cohort. For comparison the model with just CBF_2D,1PLD_ has prediction error ±12.46% [-23.00, 25.86]. Based on these factors we prefer the model CBF_3D,5PLD_ ∼ (CBF_2D,1PLD_, hb, age).

Sex is not a significant variable if added to any of the GM models in Table 3 and none of the other variables cross the critical significance *p* = 0.05 when sex is added to the models. Similarly, scanner type is not a significant variable, and adding it does not cause any other variable to cross the critical significance in either GM or WM.

### 3.3 Models of white matter CBF

Median WM CBF_2D,1PLD_ was 30.53 mL/100g/min, IQR = [23.41, 37.30]. Median WM CBF_3D,5PLD_ was 30.06 mL/100g/min [24.69, 39.49] (Figure 1(c), Table 2), not significantly different (*p* = 0.07). A Bland-Altman plot (Figure 1(d)) highlights that the fixed bias between WM CBF_3D,5PLD_ and WM CBF_2D,1PLD_ is not statistically significant. The bias is 4±17%, CI = [-29, 37%]. Nor is there significant proportional bias (*p* = 0.82, slope = 0.05 ΔCBF% / CBF_mean_, CI = [-0.41, 0.52%]). WM CBF is lower than GM CBF, implying it has a lower signal-to-noise ratio, which would increase its variability. The standard deviation of the bias between the two WM CBF techniques is wider than for GM (17% vs. 12%). Lower signal-to-noise ratio and poorer test-retest consistency in WM has been observed in many PCASL CBF studies [26–29]. Therefore, we fit models to the WM data using the same variables as for the GM data. Unlike in GM, age was not significant in WM modeling, and the relationship between WM CBF and hb, while significant, has a higher *p* value. We found the following model to predict voxel-wise mean CBF_3D,5PLD_ from CBF_2D,1PLD_, in WM:

*CBF_3D,5PLD_(WM) = 0.951×CBF_2D,1PLD_(WM) + 3.06 [mL/100g/min],* with R^2^_adj._ = 0.77, error = ±17.56% [-24.74, 44.08], and *p* < 0.001 (Figure 1(b), Table 3). Adding hb and age to the model we found:

*CBF_3D,5PLD_(WM) = 1.02×CBF_2D,1PLD_(WM) + 0.87×hb +0.08*age – 7.24* [mL/100g/min], with R^2^_adj._ = 0.78 and error = ±17.15% [-28.17, 39.06] (Figure 2(c, d)).

### 3.4 CBF relationships with other factors

In our analysis of ATT and 2D-EPI PLD_effective_, the percentage of voxels where ATT > PLD_effective_ in any scan is small: (WM: median 0.5%, IQR [0.1, 1.5], GM: median 0.1%, IQR [0.0, 0.6], max 2.9%, Figure 3). It appears that when ATT > PLD does occur, it is most commonly in the deep white matter, which could correspond to watershed regions where ATT is expected to be larger. We also found that in a multiple linear regression model ATT ∼ (hb, age), both independent variables were significant (GM: hb *p* = 0.03, age *p* < 0.001, R^2^_adj._ = 0.29; WM: hb *p* = 0.02, age *p* = 0.04, R^2^_adj._ = 0.18). In univariate analysis, ATT is significantly correlated with GM CBF_2D,1PLD_, but not with GM CBF_3D,5PLD_, WM CBF_2D,1PLD_, or WM CBF_3D,5PL_ (Figure 4). In Figure 4(a), while the trend line for GM CBF_2D,1PLD_ is not significantly non-zero, it intersects with the trend line for GM CBF_3D,5PLD_ close to 1.5 s – the PLD of the 2D-EPI 1PLD sequence – reflecting that in the ATT range we sample, the closer ATT is to PLD, the closer the two techniques are to agreement.

**Figure 3.**
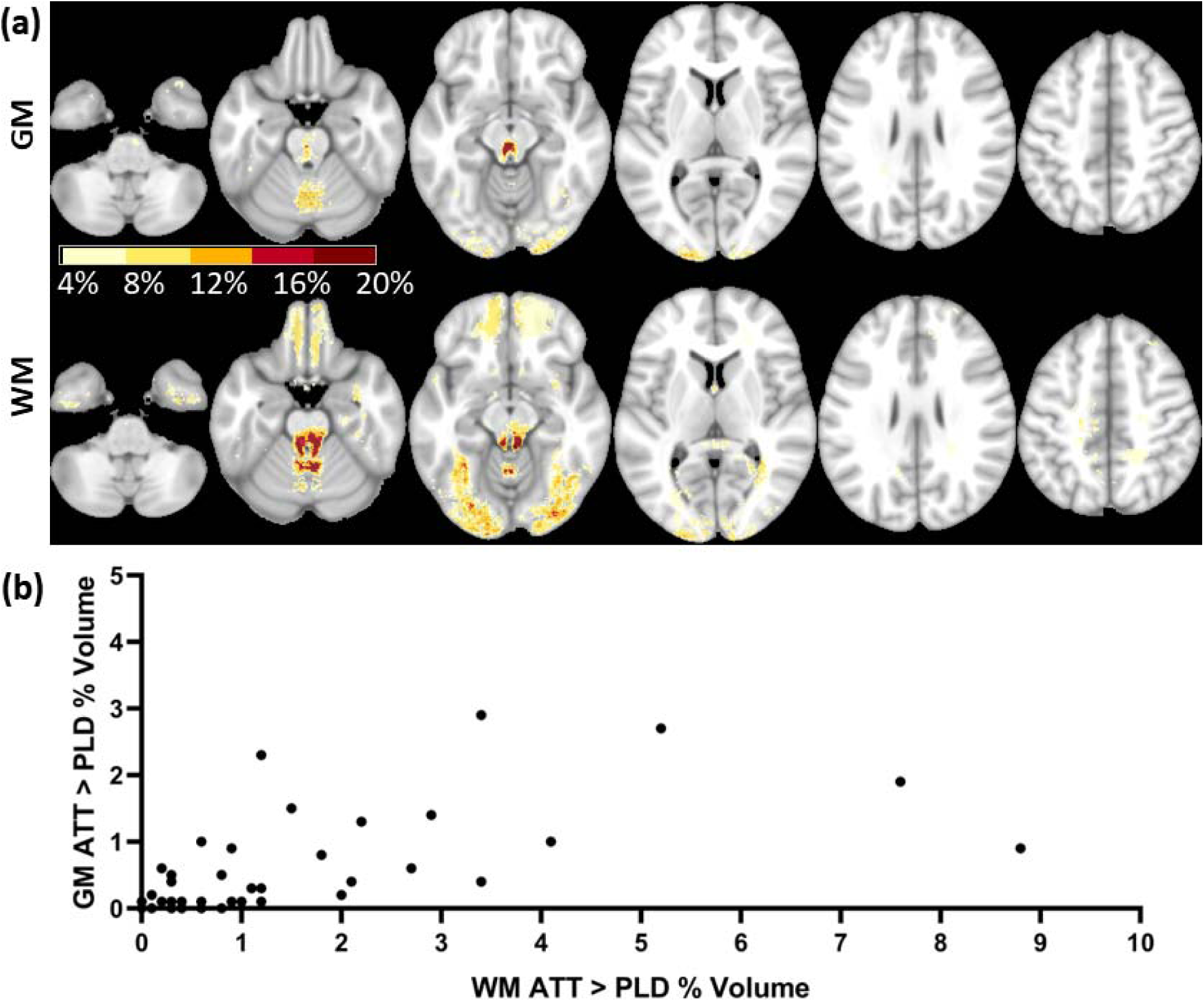
(a) Voxel-map of percentage of 50 scans with ATT > PLD, heatmap overlayed on MNI ICBM 152 atlas, 4% lower threshold for display. Montage of axial slices taken from inferior (left) to superior (right), with GM (top) and WM (bottom). (b) Scatter showing percentage of GM and WM volume per participant where ATT > PLD.

**Figure 4.**
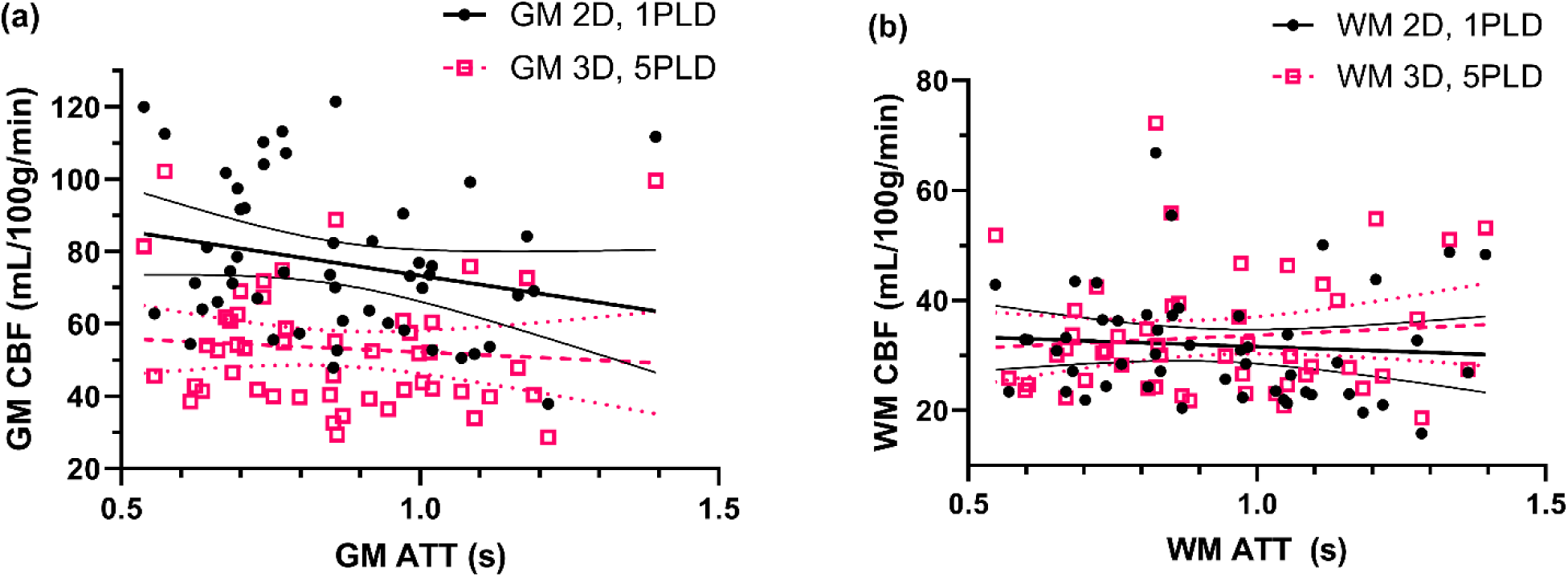
CBF vs. ATT, including scatter plots with trend lines (all nonsignificant) and 95% confidence intervals. (a) GM CBF vs. ATT (2D,1PLD: ρ = -0.26, *p* = 0.07; 3D,5PLD: ρ = -0.11, *p* = 0.46). (b) WM CBF vs. ATT.

## 4. Discussion

### 4.1 Usefulness of CBF imputation model

This study reports a simple approach for harmonization of CBF data sets collected using different MRI PCASL sequences and processing. For a cohort of 50 paired data with an expected wide range of CBF based on the age range (9–45 years) and anemia, we found linear regression models to impute 3D-GRASE 5PLD CBF from measured 2D-EPI 1PLD CBF and characterized prediction error. Models with just CBF_2D,1PLD_ or adding hb and age bothperformed well, reflecting that imputation does not require all components, but may be optimized based on available data. While it is possible that pathology or CBF-affecting features in other populations could alter the relationship between CBF_3D,5PLD_ and CBF_2D,1PLD_, our inclusion of not only healthy individuals, but also those with CBF-altering anemia, across a wide age span suggests our approach should be applicable to a variety of physiologies influencing CBF.

### 4.2 Imputation model performs similarly to acquiring another CBF scan

For the wsCV of our model including age and hb, we found 9.1% CBF_3D,5PLD_ GM variation and 11.3% WM variation between measured CBF_3D,5PLD_ values and values imputed from the model. This result is similar to previously reported values in studies examining the test-retest reliability of PCASL CBF, including wsCVs of 8.5% for GM and 12.5% for WM for 1 week test-retest of the same PCASL sequence by Chen et al., 2011 [1]. Not surprisingly, the imputation variation is slightly higher than scans collected with less intervening time. Other studies with CBF wsCV include a 1-hour test-retest with removal from the scanner between tests (5.5% for GM and 9.5% for WM), or without removal from the scanner (3.5% in GM and 8.0% in WM) [1], or with different labeling duration (6% GM and WM) [2] or alternating only radiofrequency ASL (4.8%) [30] within the same day. In terms of ICC, we calculate 0.89 for GM and 0.87 for WM imputation compared to measured, which is slightly worse than test-retest correlation found by Pfefferbaum et al., 2010 [31], who found an ICC of 0.96 with 3–7 days between scans, and Xu et al, 2010 [28], who found ICCs of 0.93 for GM and 0.96 for WM in 2–3 scans of young subjects separated by a few hours. However, imputation variability is slightly less than scans with more separation of time, as Melzer et al. reported a 13.4% wsCV found with a 13 week test-retest gap [32].

The comparability of imputing CBF_3D,5PLD_ from CBF_2D,1PLD_, age, and hb, to collecting another CBF_3D,5PLD_ measurement a few days later, indicates that imputation data can be added to a dataset to increase statistical power, with the expectation that the final dataset will be statistically similar to using directly measured data. Imputed CBF_3D,5PLD_ data points can be used in longitudinal analyses with the expectation of similar correlation as with directly measured data. Furthermore, this comparison to test-retest results suggests that, for spatially averaged CBF, using an imputation model may be preferable to rescanning a participant using a different PCASL CBF technique, even before considering the advantage of no additional effort and expense once the paired data for an imputation model is acquired. We had the advantage that our data set was primarily from one scanner, and the 8% (4 scans) from another scanner were from the same scanner model (Siemens, Prisma). A study of variability in PCASL CBF across different scanner models shows significant fixed bias, but results co-vary [3], so in a study with multiple scanner models, scanner effect may be effectively corrected for by adding it as a variable in the model. Our results support the feasibility and utility of harmonizing other datasets with CBF by PCASL techniques.

### 4.3 CBF_2D,1PLD_ vs. CBF_3D,5PLD_ differences

Based on the Bland-Altman calculation of bias (GM bias -37%, CI = [-61, -13] %; WM bias -4%, CI = [-29, 17] %), most of the bias between the two techniques, even applied to varied populations including those with sickle cell disease and children, may be explained by one or more proportional factors affecting the GM.

Unlike for CBF_3D,5PLD_, ATT is not solved for and included in the calculation of CBF_2D,1PLD_, so measurement variance due to differences between ATT and PLD could bias CBF_2D,1PLD_ compared to the CBF_3D,5PLD_ technique. CBF_2D,1PLD_ is significantly anti-correlated with ATT in the GM, whereas GM CBF_3D,5PLD_ is not (Figure 4). In WM, CBF_2D,1PLD_ is not significantly related to ATT, and has a much smaller bias compared to WM CBF_3D,5PLD_. The closer ATT is to the PLD of our 2D PCASL CBF technique, the more likely the techniques are to agree (Figure 4). An extreme case of ATT artifact is when ATT > PLD, i.e., the labeled blood has yet to arrive at a voxel. We find this condition is true for only a small fraction of voxels in any scan in our cohort (Figure 3).

The ATT may also influence labeling efficiency, another potential source of error in CBF calculation [33]. Both techniques should be affected to the same proportion by errors in labeling efficiency, so this should not be a source of bias. However, labeling efficiency and ATT should vary inversely. At high blood velocity, ATT will be shorter, and blood will not remain in the labeling plane for as long, decreasing labeling efficiency. Age and hb significantly correlate with ATT, so they may be acting as proxy variables predicting ATT bias in CBF_2D,1PLD_, which could explain why they are more significant in the GM model than in the WM model.

Our CBF data were calculated using a one-compartment model, which is most recommended for research. More complex modeling may improve technique agreement prior to any post-hoc modelling of corrections, for example, by taking into account different susceptibility decay rates of magnetically labeled water in the blood vs. after perfusion into the tissue compartment. Such decay rate differences may affect PCASL data differently for different PLDs, thus creating a bias between two techniques with different PLDs. However, in the case of our data, a single-compartment model is, sufficient to enable reuse of our lower resolution, 2D readout PCASL CBF data with higher resolution, 3D readout data, which is in agreement with consensus recommendations that decay rate differences may be safely ignored in the appropriate research applications [5].

### 4.4 Limitations

Although our cohort and subgroup sizes are comparable to similar studies, our study is still subject to the effects of small N for statistical approximations [4]. However, because we use paired data in our study, this is only a limitation if there are non-representative features of our cohort that also affect the relationship between CBF_2D,1PLD_ and CBF_3D,5PLD_, and we have intentionally selected our cohort to account for known possible factors.

We studied a wider range of hb by combining participants with and without SCD. We cannot fully differentiate between effects of hb and other effects of SCD.

Another limitation is that we discuss models only for spatially averaged, partial-volume corrected GM and WM ROIs. Imputation of CBF for small subregions may not be as reliable and would require additional studies.

### 4.5 Conclusion

Using 50 paired datapoints, we created linear regression models to predict CBF from 3D-GRASE 5PLD PCASL using 2D-EPI 1PLD PCASL, applicable to a range of hb and age values. The performance of these models was similar to the test-retest performance of CBF from PCASL scans collected a few days apart. Much of the discrepancy between the two techniques may be accounted for by unidentified ATT artifacts on CBF_2D,1PLD_ that are corrected for in CBF_3D,5PLD_ by multi-PLD modeling of ATT, and the variability in the CBF_2D,1PLD_ and CBF_3D,5PLD_ techniques. This or similar harmonizations should enable extant, effort-intensive PCASL CBF datasets to be reused alongside datasets generated by different PCASL CBF techniques.

## Data Availability

All data produced in the present study are available upon reasonable request to the authors.

## Acknowledgements

We would like to acknowledge Jade Beshears, Maria Hagan, Nkemdilim Igwe, and Heather Quartuccio for contributions to study coordination.

Statistical analysis was generated using SAS software, Version 9 of the SAS System for Linux. Copyright © 2012–2020 SAS Institute Inc. SAS and all other SAS Institute Inc. product or service names are registered trademarks or trademarks of SAS Institute Inc., Cary, NC, USA. Additional statistical analysis was conducted using scikitlearn [34] and statsmodels [35] libraries for the Python programming language.

Research reported in this publication was supported by the Washington University Institute of Clinical and Translational Sciences grant UL1TR002345 from the National Center for Advancing Translational Sciences (NCATS); the Eunice Kennedy Shriver National Institute Of Child Health & Human Development of the National Institutes of Health (NIH) under Award Number P50 HD103525 to the Intellectual and Developmental Disabilities Research Center at Washington University; by the NIH National Institute of Neurological Disorders and Stroke (K23NS099472 [KPG] and R01NS121065 [KPG], RF1NS116565 [HA, ALF], R21NS127425 [HA]); and by the NIH National Heart, Lung, and Blood Institute (R01HL157188 [MEF] and R01HL129241 [ALF, HA]). The content is solely the responsibility of the authors and does not necessarily represent the official views of the National Institutes of Health.

The data that support the findings of this study are available from the corresponding author upon reasonable request.

## Author Contributions

**Josiah B. Lewis:** Conceptualization, Methodology, Software, Formal Analysis, Investigation, Data Curation, Writing – Original Draft, Visualization. **Chunwei Ying:** Validation, Software, Writing – Review & Editing. **Michael M. Binkley**: Formal Analysis, Writing – Review & Editing. **Melanie E. Fields**: Writing – Review & Editing, Funding acquisition. **Igor Dedkov:** Investigation, Software, Writing – Review & Editing, Visualization. **Slim Fellah:** Validation, Software, Investigation, Writing – Review & Editing. **Jingyi Zhang:** Formal Analysis, Writing – Review & Editing. **Amy Mirro**: Software, Writing – Review & Editing. **Joshua S. Shimony**: Writing – Review & Editing. **Yasheng Chen**: Writing – Review & Editing. **Jin-Moo Lee**: Writing – Review & Editing. **Andria L. Ford**: Writing – Review & Editing, Funding acquisition. **Hongyu An**: Writing – Review & Editing, Funding acquisition. **Kristin P. Guilliams**: Conceptualization, Methodology, Investigation, Writing – Review & Editing, Supervision, Funding acquisition.

## Appendices

**Figure A. 1.**
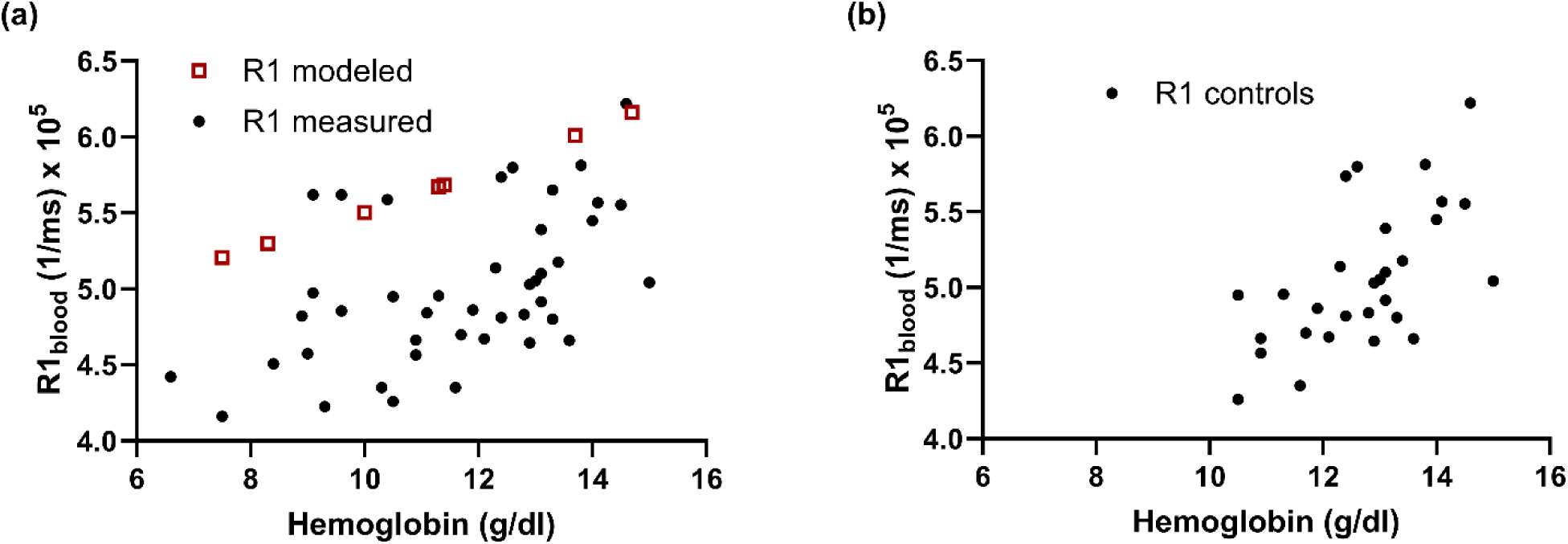
R1blood relaxation rates (T1blood=1/R1blood) used in this study by participant, either measured from the fast inversion recovery signal inside a hand-drawn region of interest in the superior sagittal sinus (filled black circles) or modeled based on the hemoglobin measurement (open red squares). (a) shows all measurements, while (b) shows that R1s from the control subgroup have a more linear with hemoglobin (i.e., with hemoglobin A).

## Appendix A

We calculated the confidence interval for our models using linear interpolation between points in the array of leave-one-out cross-validation errors, and calculated the prediction error (standard deviation) from the upper and lower limits of the confidence interval, *u* and *l*: error = ± (u - l) / (2 × 1.96).

We calculated the within-subject coefficient of variation, wsCV, as follows:

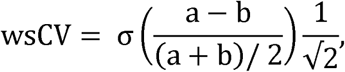

that is, 1/√2 times the standard deviation of the test-retest difference between measures a and b, where test-retest difference is calculated as a fraction of the mean.

We calculated the interclass correlation coefficient (ICC) as follows:

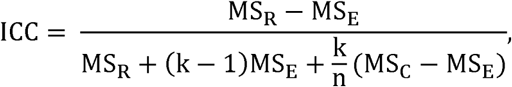

where *MS* for *R*, *C*, and *E* are the mean squared terms for row (intrasubject), column (intratest), and error (residual) effects, respectively. *k* is the number of pseudo-tests (measured and imputed CBF_3D,5PLD,_ so *k* = 2), and *n* is the number of participants.

Using ANOVA we tested pairs of nested models with the null hypothesis that the simpler model with *RSS_1_* and degrees of freedom *DoF_1_*performs no worse than the model with more terms with *RSS_2_* and *DoF_2_*. The *p* value for the null hypothesis was calculated from the *F*-distribution for degrees of freedom *DoF_1_*– *DoF_2_* and *DoF_2_*, evaluated at:

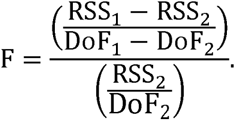

We tested whether the reported model comparisons would cross critical significance (*p* = 0.05) after correction for multiple comparisons. Because the models shared some independent variables, and to balance control of type I and type II error rates, we used the Benjamini-Hochberg method. We reported unadjusted *p* values for clarity.

