## Supplementary Material for "Harmonization of cerebral blood flow measurements by multi-delay 3D gradient and spin echo, and single-delay 2D echo planar imaging"

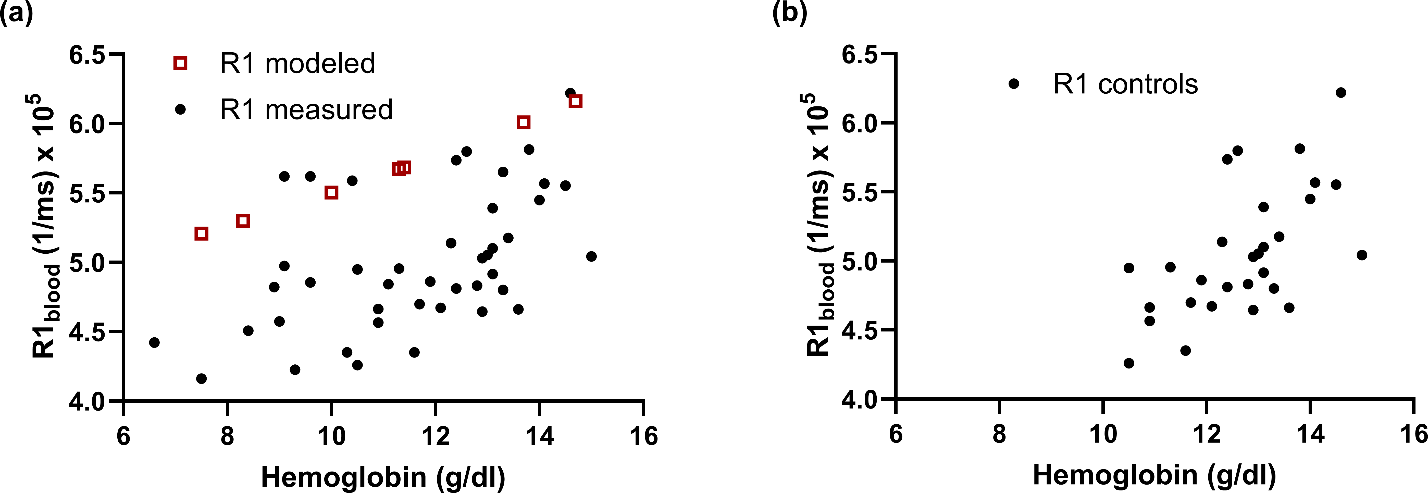

**Figure S1.** R1_blood_ relaxation rates (T1_blood_=1/R1_blood_) used in this study by participant, either measured from the fast inversion recovery signal inside a hand-drawn region of interest in the superior sagittal sinus (filled black circles) or modeled based on the hemoglobin measurement (open red squares). (a) shows all measurements, while (b) shows that R1s from the control subgroup have a more linear with hemoglobin (i.e., with hemoglobin A).

**Supplementary Methods**

We calculated the confidence interval for our models using linear interpolation between points in the array of leave-one-out cross-validation errors, and calculated the prediction error (standard deviation) from the upper and lower limits of the confidence interval, *u* and *l*: error = ± (u - l) / (2 × 1.96).

We calculated the within-subject coefficient of variation, wsCV, as follows:

$$wsCV= \sigma\left( \frac{a-b}{\left( a+b \right)/ 2} \right)\frac{1}{\surd2},$$

that is, $1/\surd2$ times the standard deviation of the test-retest difference between measures a and b, where test-retest difference is calculated as a fraction of the mean.

We calculated the interclass correlation coefficient (ICC) as follows:

$$ICC= \frac{MS_{R}-MS_{E}}{MS_{R}+\left( k-1 \right)MS_{E}+\frac{k}{n}(MS_{C}-MS_{E})},$$

where *MS* for *R*, *C*, and *E* are the mean squared terms for row (intrasubject), column (intratest), and error (residual) effects, respectively. *k* is the number of pseudo-tests (measured and imputed CBF_3D,5PLD,_ so *k* = 2), and *n* is the number of participants.

Using ANOVA we tested pairs of nested models with the null hypothesis that the simpler model with *RSS_1_* and degrees of freedom *DoF_1_* performs no worse than the model with more terms with *RSS_2_* and *DoF_2_*. The *p* value for the null hypothesis was calculated from the *F*-distribution for degrees of freedom *DoF_1_* – *DoF_2_* and *DoF_2_*, evaluated at:

$$F=\frac{\left( \frac{\mathrm{RS}S_{1}-RSS_{2}}{\mathrm{Do}F_{1}-DoF_{2}} \right)}{\left( \frac{\mathrm{RS}S_{2}}{\mathrm{Do}F_{2}} \right)}.$$

We tested whether the reported model comparisons would cross critical significance (*p* = 0.05) after correction for multiple comparisons. Because the models shared some independent variables, and to balance control of type I and type II error rates, we used the Benjamini-Hochberg method. We reported unadjusted *p* values for clarity.
